# Awareness, treatment, and control of diabetes in Bangladesh: evidence from the Bangladesh Demographic and Health Survey 2017/18

**DOI:** 10.1101/2021.07.09.21260274

**Authors:** Md. Nuruzzaman Khan, John C. Oldroyd, Mohammad Bellal Hossain, Rakibul M. Islam

**Author notes:** Corresponding authors: Md Nuruzzaman Khan, Lecturer, Department of Population Science, Jatiya Kabi Kazi Nazrul Islam University, Mymensingh, Bangladesh.

## Abstract

**Background:** The prevalence of diabetes is increasing in Bangladesh; however, the management of diabetes assessed by diabetes awareness, treatment, and control, remains poor. We aimed to estimate the age-standardised prevalence of awareness, treatment, and control of diabetes and its associated factors.

**Methods:** In this cross-sectional study, data from 1,174 Bangladeshi adults aged 18 years and older available from the most recent nationally representative Bangladesh Demographic and Health Survey (BDHS) 2017-18 were analysed. Awareness, treatment, and control of diabetes were our outcomes of interest. Age-stadarised prevalence of awareness, treatment, and control were estimated using the direct standardisation. Multilevel mixed-effects Poisson regression models were used to identify factors associated with awareness, treatment, and control of diabetes.

**Results:** Among those with diabetes (n=1,174), only 30.9% (95% CI, 28.2-33.6) were aware that they had the condition, and 28.2% (95% CI, 25.6-30.7) were on treatment. Among those treated for diabetes, only 26.5% (95% CI, 19.5-33.5) had controlled diabetes. Prevalences for awareness, treatment and control were even lower in men than women. Factors positively associated with awareness and treatment were increasing age and hypertension while factors negatively associated with awareness and treatment were being men and having lower education. Factors associated with poor control were secondary education and residing in Rajshahi and Rangpur divisions.

**Conclusion:** This study provides evidence of poor management of diabetes in Bangladesh, especially in men. Less than one-third of the people with diabetes were aware of their condition. Just over one-fourth of the people with diabetes were on treatment, and those were treated one-fourth had controlled diabetes. Interventions targeting younger people, in particular men and those with lower education, are urgently needed. The government needs to strengthen diabetes management programs within primary health care and address structural factors including the costs of diabetes care to improve awareness, treatment, and control of diabetes in Bangladesh.

## Background

Globally, the prevalence of diabetes mellitus is steadily increasing [1]. Currently, ∼463 million people worldwide have diabetes, and a further 374 million people are at increased risk of developing diabetes [2] which is expected to reach 700 million by 2045 [3]. The majority of this increase will occur in low- and lower-middle-income countries (LMICs), particularly in Asia and Africa [3]. The epidemiological transition leads to such rapid increases in diabetes prevalence in LMICs mainly due to the westernisation of diets, reduced physical activity, changing patterns in leisure activities, longer working hours, and reduced sleep [4].

Diabetes mellitus causes ∼1.6 million deaths annually worldwide [2, 5]. The majority of these deaths are premature, occurring in younger age groups [2, 3]. Moreover, diabetes management is associated with some 12% of the global health expenditure [5]. The Sustainable Development Goals (SDGs) have set a target to reduce premature death from non-communicable diseases (NCDs) by one-third by 2030 as compared to 2015 levels (SDG Target 3.4) [6]. The effective management of diabetes (awareness, treatment, and control) can reduce diabetes-associated complications, deaths, and costs, and can help to achieve the SDGs’ target [7, 8].

Bangladesh has one of the highest burdens of diabetes among countries in the Southeast Asian region (∼25 million cases) with evidence this disease burden is increasing [9]. Though the prevalence of diabetes is increasing in Bangladesh, there is a lack of healthcare facilities for diabetes management, especially in rural areas [10, 11]. The majority of facilities are private urban facilities [12], which are expensive [13]. Therefore, many people cannot go for diabetes diagnosis and/or afford diabetes treatment, which increases diabetes complications and deaths [13].

Several studies have reported a low level of diabetes management in Bangladesh [11, 14-16]. However, these studies are limited by small sample size, a selected population (e.g., specific age band) and/or reported unstandardised estimates. To our knowledge, the age-standardised prevalences of awareness, treatment, and control of diabetes; and factors associated with these conditions among Bangladeshi adults 18 years and older using the latest Bangladesh Demographic and Health Survey (BDHS) 2017-18 data have not yet been estimated. Therefore, we aimed to determine the prevalence of these conditions and associated factors in the Bangladeshi adult population.

## Methods

### Data

The data used were extracted from the latest BDHS 2017/18 [17]. This survey is nationally representative and conducted between October 24, 2017 and March 15, 2018 by the National Institute of Population Research and Training under the Ministry of Health and Family Welfare of Bangladesh and Mitra and Associates. The survey samples were selected using a two-stage stratified random sampling method. At the first stage, 675 primary sampling units (PSUs) were selected randomly from the list of enumeration areas generated as part of the Bangladesh Population and Housing Census 2011. Three PSUs were excluded from the survey due to flood. In the second stage, 30 households were selected from each PSU through probability proportional to the size. This process produced a list of the 19,584 eligible households, and interviews were completed in 19,457 households (overall household response rate 99.4%) [17]. Data associated with NCDs, including diabetes and hypertension were collected from one in every four included households. The detailed sampling procedure has been published elsewhere [17]. For comparison purposes, the BDHS 2011 data were also analysed, however, this survey collected NCDs data from respondents aged 35 and older only [18].

### Outcomes measures

Awareness, treatment, and control of diabetes were our outcome variables. These outcome variables were generated for the respondents who had diabetes which was defined as having elevated fasting blood glucose (FBG, ≥7 mmol/L) and/or on blood glucose lowering medication at the time of the survey [9]. Awareness of diabetes refers to respondents reported knowing their glucose level as measured before and/or ever been told by a doctor or nurse that they have diabetes. Treatment of diabetes refers to respondents using medication to control their diabetes. Control of diabetes refers to treated diabetes with FBG value less than 7.0 mmol/L.

### Explanatory variables

The factors associated with awareness, treatment, and control of diabetes considered in this study were selected through a comprehensive literature review in Bangladesh and its neighboring countries [9, 11, 19-22]. Factors included were respondents’ age group (18-34, 35-39, 40-44, 45-49, 50-54, 55-59, 60-64, ≥65 years), sex (men, women), education (no education/pre-primary, primary, secondary, higher), working status (currently working, yes/no), hypertension status (yes, no), wealth quintile (poorest, poorer, middle, richer, and richest), place of residence (urban/rural), and administrative division at the time of the survey. Respondents were considered hypertensive if they had (i) systolic blood pressure ≥140 mmHg and/or a diastolic blood pressure ≥90 mmHg, or (ii) taking any prescribed drugs to control blood pressure [23]. The wealth quintile was generated using principal component analysis based on the respondents’ durable and non-durable household goods, such as ownership of land, housing materials, electricity, and television.

### Statistical analysis

We used descriptive statistics to describe the individual, household, and community-level characteristics of the respondents. Age-standardized prevalence estimates of awareness, treatment, and control of diabetes were calculated and presented with 95% CI. The age-standardised estimates were by direct standardisation based on the Bangladesh Population and Housing Census 2011.

We used multilevel mixed-effects Poisson regression with a robust variance to identify factors associated with awareness, treatment, and control of diabetes mellitus. The results were presented as a prevalence ratio (PR) with 95% CI. Poisson regression was used to avoid overestimation of the odds ratios that occur using logistic regression in the cross-sectional studies when the outcome of interest is common [24]. Furthermore, in the BDHS, individuals were nested within the household; households were nested within the PSU/cluster. Therefore, our multilevel mixed-effects Poisson regression model accounts for these multiple hierarchies and dependency in data and the problem of overestimation. Tests for multi-collinearity and effect modification between explanatory variables were checked before entering into models. All statistical tests were two-sided, and a p-value <0·05 was considered statistically significant. The Strengthening the Reporting of Observational Studies in Epidemiology (STROBE) guidelines informed the design and reporting of the study [25]. All analyses used statistical software package Stata (version 15·1; Stata Corp LP, College Station, Texas).

## Results

The individual, household, and community-level characteristics of 1,174 participants who provided complete data are summarised in Table 1. The mean (standard error) age of the study participants was 46.67 (0.46) years, and 54% were women. A quarter of the participants had no formal education, 46% were employed, and 65% resided in urban areas. Twenty percent of participants were overweight/obese, and 46% were hypertensive.

**Table 1:**
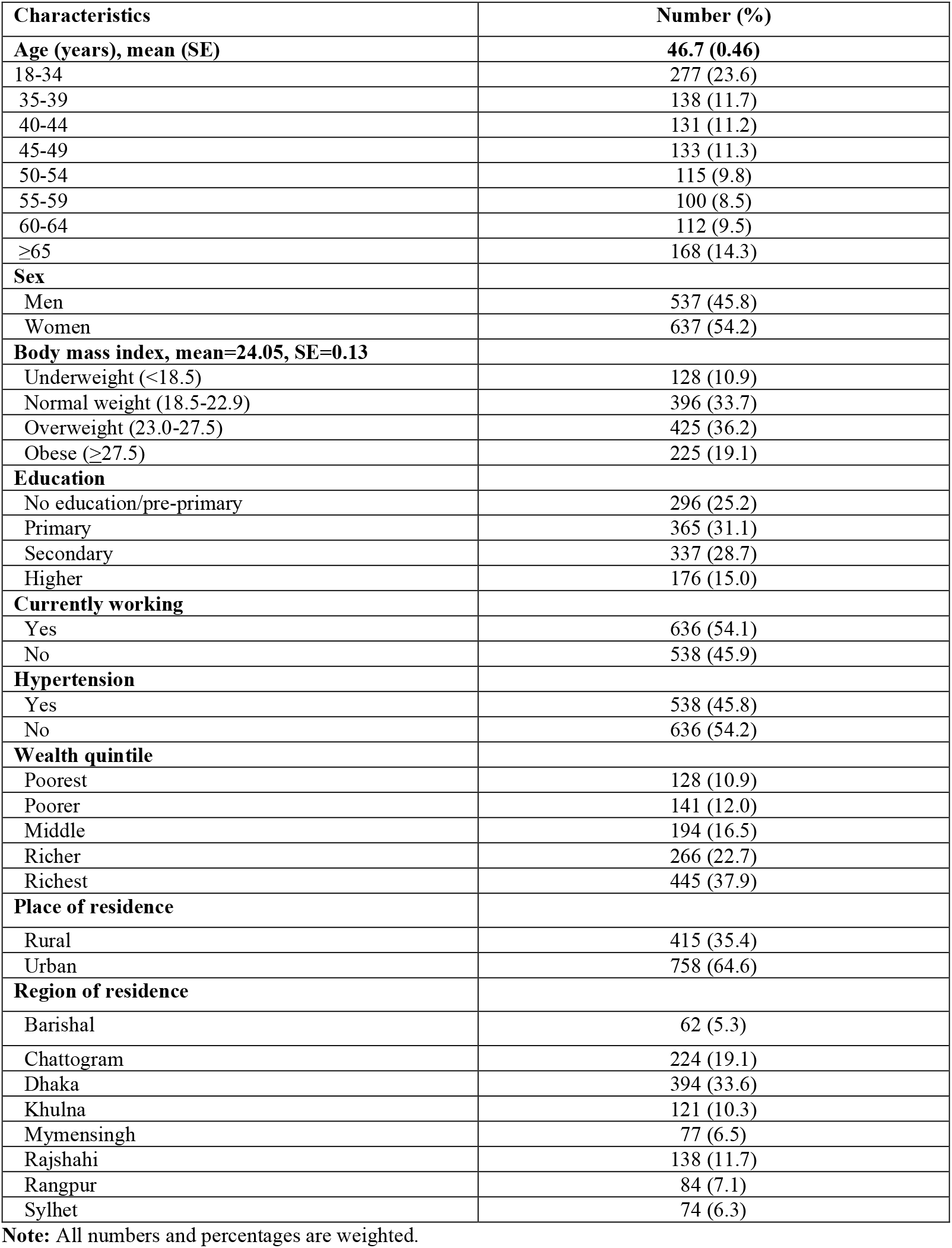
Background characteristics of the study population (N=1,174)

The age-standardised prevalence of awareness, treatment, and control of diabetes are presented in Table 2. The overall age-standardised prevalence of awareness was 30.9% (95% CI, 28.2-33.6). People ≥65 years had the highest prevalence (57.0%, 95% CI, 49.5-64.4) compared to other ages. The prevalence in men was 24.5% (95% CI, 21.1-28.0) compared to women 35.0% (95% CI, 31.3-38.7). The overall prevalence of treatment was 28.2.3% (95% CI, 25.6-30.7). Treatment was higher among women (32.1%, 95% CI, 28.5-35.7) than men (22.1%, 95% CI, 18.8-25.3). The prevalence of treatment increased with increasing age, a trend found in men and women. The prevalence of control of diabetes was 26.5% (95% CI, 19.5-33.5), increasing with increasing respondent age. The prevalence of control was higher among women (27.4%, 95% CI, 19.4-35.4) than men (23.9%, 95% CI, 5.5-39.4).

**Table 2:** Age-standardized prevalence of awareness, treatment, and control of diabetes by sex, Bangladesh 2017/18.

| Age in years | Awareness |  |  | Treatment |  |  | Control |  |  |
| --- | --- | --- | --- | --- | --- | --- | --- | --- | --- |
|  | Overall (453/1174) | Men (198/537) | Women (255/637) | Overall (413/1174) | Men (178/537) | Women (235/637) | Overall (126/413) | Men (58/178) | Women (68/235) |
| 18-34 | 15.1 (10.8-19.3) | 6.8 (1.9-11.7) | 19.9 (14.0-25.8) | 13.3 (9.3-17.3) | 5.8 (1.3-10.4) | 17.6 (12.0-23.3) | 21.6 (8.1-35.1) | 16.7 (-16-49.6) | 22.6 (7.5-37.6) |
| 35-39 | 32.9 (25.0-40.8) | 28.6 (16.6-40.5) | 35.7 (25.4-46.0) | 30.0 (22.4-37.6) | 25.0 (13.5-36.5) | 33.3 (23.2-43.5) | 26.2 (12.7-39.7) | 28.6 (3.8-53.3) | 25.0 (8.6-41.4) |
| 40-44 | 37.9 (29.3-46.5) | 25.0 (13.5-36.5) | 48.5 (36.5-60.5) | 33.1 (24.7-41.4) | 21.4 (10.6-32.3) | 42.6 (30.8-54.5) | 29.3 (15.1-43.4) | 25.0 (-1-50.8) | 31.0 (13.8-48.3) |
| 45-49 | 51.5 (42.9-60.1) | 46.7 (31.9-61.4) | 54.0 (43.4-64.6) | 47.7 (39.2-56.3) | 40.0 (25.5-54.5) | 51.7 (41.1-62.3) | 30.2 (18.7-41.6) | 33.3 (10.7-55.9) | 28.9 (15.4-42.3) |
| 50-54 | 48.6 (39.3-58.0) | 36.4 (23.5-49.2) | 60.7 (47.8-73.6) | 45.0 (35.7-54.4) | 34.5 (21.8-47.3) | 55.4 (42.2-68.5) | 30.0 (17.1-42.9) | 36.8 (14.4-59.3) | 25.8 (10.1-41.5) |
| 55-59 | 59.1 (49.9-68.3) | 64.4 (50.3-78.6) | 55.4 (43.2-67.6) | 55.4 (46.1-64.8) | 64.4 (50.3-78.6) | 49.2 (37.0-61.5) | 29.5 (17.1-42.9) | 27.6 (10.9-44.3) | 31.3 (14.8-47.6) |
| 60-64 | 55.2 (45.7-64.8) | 60.3 (47.6-73.1) | 48.9 (34.5-63.4) | 52.4 (42.8-62.0) | 56.9 (44.0-69.8) | 46.8 (32.4-61.3) | 34.5 (21.8-47.3) | 33.3 (16.9-49.8) | 36.4 (15.7-57.0) |
| ≥65 | 57.0 (49.5-64.4) | 58.2 (48.3-68.0) | 55.4 (44.0-66.8) | 52.9 (45.4-60.4) | 52.0 (42.1-62.0) | 54.1 (42.6-65.5) | 41.8 (21.8-47.3) | 35.3 (21.9-48.6) | 50.0 (34.2-65.8) |
| Total | 30.9 (28.2-33.6) | 24.5 (21.1-28.0) | 35.0 (31.3-38.7) | 28.2 (25.6-30.7) | 22.1 (18.8-25.3) | 32.1 (28.5-35.7) | 26.5 (19.5-33.5) | 23.9 (8.5-39.4) | 27.4 (19.4-35.4) |

**Note:** Awareness of diabetes refers that the respondents ever been told by a doctor or nurse that they have diabetes; Treatment of diabetes refers to respondents using medication to control their diabetes; Control of diabetes refers to treated diabetes with FBG value less than 7.0 mmol/L.

Figure 1 shows comparisons of diabetes prevalence and diabetes awareness, treatment, and control between 2011 and 2017/18 BDHSs’ surveys in respondents aged 35 and older. We observed a significant increase in diabetes in Bangladesh between 2011 and 2017/18, from 10.9% to 13.7% (*p*<0.001 for difference). In 2011, 35% of the people with diabetes reported that they were aware of their condition, compared with 46% in 2017/18. A third of the people with diabetes were treated in 2011 compared with 42% in 2017/18. However, the proportion of controlled diabetes declined between the surveys, from 36.4% in 2017/18 to 31.4% in 2011.

**Figure 1:**
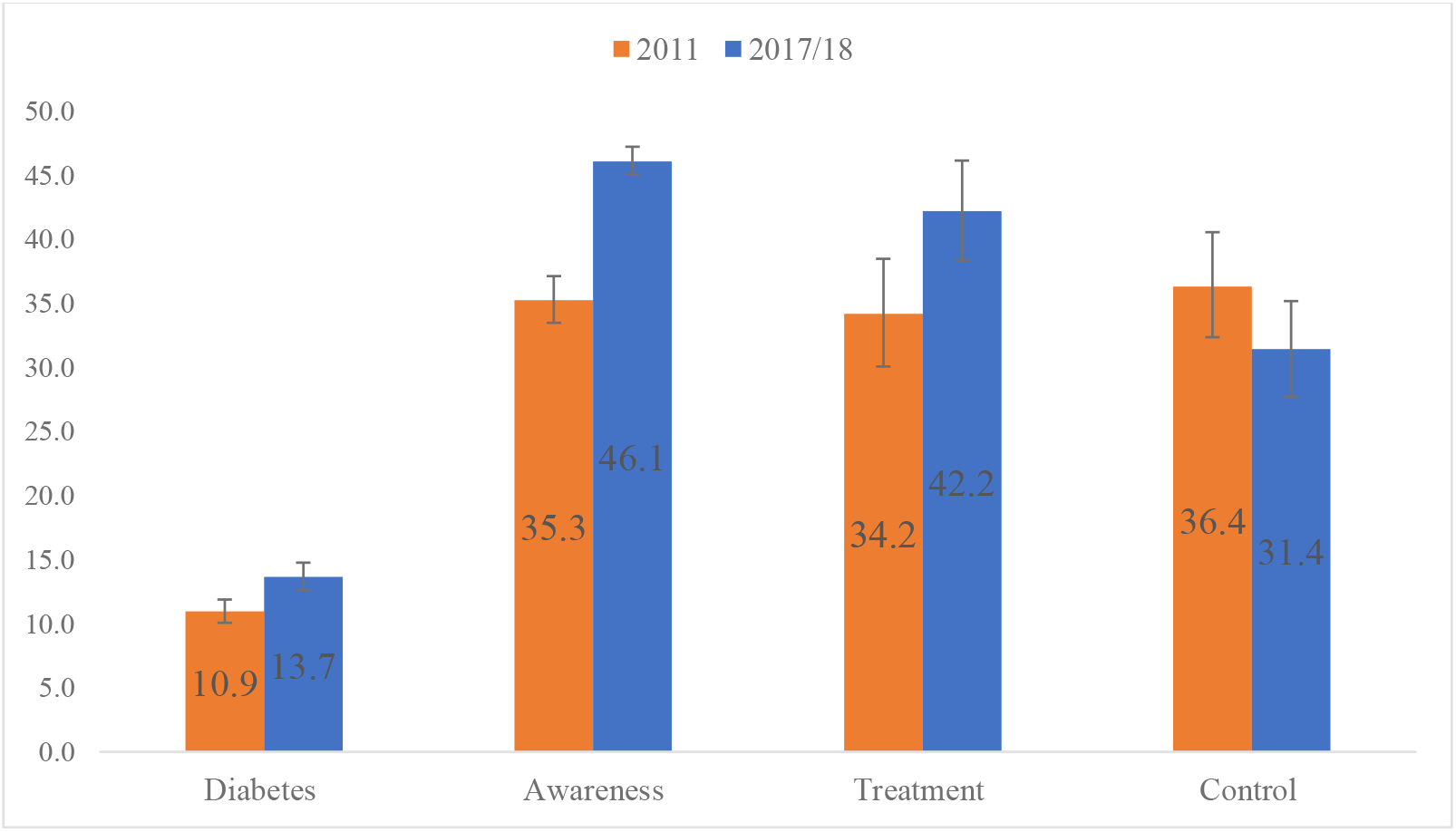
Comparisons of diabetes and its management between 2011 and 2017/18 BDHSs’ surveys. **Note:** The 2011 BDHS collected data from the respondents aged 35 and over, whereas 2017/18 BDHS collected data from the respondents aged 18 and over. For comparison, data presented are in respondents aged 35 and over only.

Factors associated with awareness, treatment, and control of diabetes are presented in Table 3. A gradual increase of awareness with increasing respondents’ age, was observed. Women were more likely to be aware of diabetes than men (PR, 1.34, 95% CI, 1.08-1.65). Underweight respondents were less likely than normal-weight respondents to be aware of diabetes (PR, 0.61, 95% CI, 0.40-0.93). Respondents with primary, pre-primary or no education were less likely to be aware (PR, 0.74, 95% CI, 0.59-0.93) than respondents with higher education. The respondents who belonged to the higher wealth quintile were more likely to be aware (PR, 1.41, 95% CI, 1.00-1.99) than the lower wealth quintile. Those with hypertension were more likely to be aware (PR 1.39 (95% CI 1.17-1.64) than those without hypertension. Awareness of diabetes was less likely among respondents residing in the Dhaka division (PR, 0.70, 95% CI 0.52-0.95) than those residing in the Barishal division.

**Table 3:** Factors associated with the awareness, treatment, and control of diabetes in Bangladesh, 2017/18.

|  | **Awareness**<br>**PR (95% CI)** | **Treatment**<br>**PR (95% CI)** | **Control**<br>**PR (95% CI)** |
| --- | --- | --- | --- |
| **Individual level** |  |  |  |
| *Age in years (ref: 18-34)* |  |  |  |
| 35-39 | 2.12 (1.46-3.07)*** | 2.30 (1.53-3.44)*** | 0.95 (0.43-2.12) |
| 40-44 | 2.45 (1.65-3.64)*** | 2.49 (1.61-3.84)*** | 1.37 (0.63-2.96) |
| 45-49 | 3.11 (2.19-4.43)*** | 3.21 (2.19-4.71)*** | 1.93 (0.93-4.03) |
| 50-54 | 3.10 (2.14-4.48)*** | 3.22 (2.15-4.82)*** | 1.21 (0.52-2.79) |
| 55-59 | 3.44 (2.42-4.91)*** | 3.69 (2.49-5.45)*** | 1.23 (0.57-2.65) |
| 60-64 | 3.65 (2.53-5.27)*** | 3.88 (2.62-3.75)*** | 1.75 (0.83-3.69) |
| ≥65 | 3.70 (2.63-5.21)*** | 3.72 (2.54-5.45)*** | 1.92 (0.97-3.81) |
| *Sex (ref: men)* | 1.34 (1.08-1.65)*** | 1.32 (1.06-1.64)*** | 1.04 (0.69-1.56) |
| *Body Mass Index (kg/m<sup>2</sup>) (ref: normal weight)* |  |  |  |
| Underweight (<18.5) | 0.61 (0.40-0.93)** | 0.68 (0.44-1.04) | 1.68 (0.99-2.86) |
| Overweight (23.0-27.5) | 0.98 (0.82-1.18) | 0.97 (0.80-1.19) | 0.79 (0.55-1.14) |
| Obese (≥27.5) | 1.01 (0.81-1.26) | 0.99 (0.78-1.26) | 1.09 (0.70-1.70) |
| **Education (ref: higher education)** |  |  |  |
| No education/pre-primary/primary | 0.74 (0.59-0.93)*** | 0.69 (0.54-0.89)*** | 0.81 (0.54-1.23) |
| Secondary | 1.03 (0.83-1.28) | 0.97 (0.76-1.23) | 0.49 (0.29-0.83)*** |
| *Currently working (ref: no)* | 1.07 (0.89-1.29) | 1.02 (0.84-1.24) | 1.31 (0.90-1.92) |
| *Hypertension (ref: no)* | 1.39 (1.17-1.64)*** | 1.44 (1.19-1.74)*** | 0.91 (0.68-1.22) |
| **Household level** |  |  |  |
| *Wealth quintile (ref: lowest)* |  |  |  |
| Second | 0.80 (0.55-1.18) | 0.72 (0.48-1.08) | 0.68 (0.35-1.31) |
| Middle | 1.11 (0.78-1.58) | 0.97 (0.67-1.41) | 1.02 (0.64-1.61) |
| Fourth | 1.17 (0.83-1.64) | 1.07 (0.75-1.53) | 0.63 (0.36-1.11) |
| Highest | 1.41 (1.00-1.99)** | 1.27 (0.87-1.82) | 0.65 (0.40-1.06) |
| **Community level** |  |  |  |
| *Place of residence (ref: rural)* | 0.91 (0.77-1.07) | 0.94 (0.79-1.11) | 1.06 (0.76-1.47) |
| *Administrative division (ref: Barishal)* |  |  |  |
| Chattogram | 1.01 (0.78-1.32) | 1.03 (0.77-1.35) | 0.75 (0.44-1.27) |
| Dhaka | 0.70 (0.52-0.95) | 0.68 (0.50-0.93) | 0.60 (0.34-1.04) |
| Khulna | 0.97 (0.73-1.28) | 1.00 (0.76-1.34) | 0.73 (0.41-1.29) |
| Mymensingh | 0.97 (0.71-1.33) | 0.99 (0.71-1.39) | 0.95 (0.54-1.70) |
| Rajshahi | 1.19 (0.90-1.58) | 1.09 (0.81-1.46) | 0.48 (0.27-0.86)*** |
| Rangpur | 1.19 (0.88-1.60) | 1.28 (0.94-1.73) | 0.53 (0.29-0.98)** |
| Sylhet | 0.86 (0.63-1.16) | 0.90 (0.66-1.21) | 1.04 (0.61-1.77) |

PR = prevalence ratio, \*\*\*p<0.01, \*\*p<0.05

We found the likelihood of treatment of diabetes increased with the increase of respondents’ age. The highest likelihood of treatment was in respondents aged 60-64 years (PR 3.88, 95% CI, 2.62-3.75) compared to the respondents aged 18-34 years. Women were more likely to be treated for diabetes than men (PR, 1.32, 95% CI, 1.06-1.64). Respondents with primary, pre-primary or no education were less likely to be treated than the respondents with higher education (PR 0.69, 95% CI, 0.54-0.89). Respondents with hypertension were more likely to be treated for diabetes than the respondents without hypertension (PR 1.44 95% CI, 1.19-1.74).

The likelihood of control of diabetes was found to be 51% (PR, 0.49, 95% CI, 0.28-0.83) lower among secondary educated respondents than the higher educated respondents. We found a lower likelihood of control of diabetes among respondents in Rajshahi (PR 0.48, 95% CI, 0.27-0.86) and Rangpur (PR 0.53, 95% CI, 0.29-0.98) division as compared to residents in the Barishal division.

## Discussion

In this nationally representative study of Bangladesh using data from the most recent BDHS 2017-18, we estimated the age-standardised prevalence of awareness, treatment, and control of diabetes and identified factors associated with these conditions. Our findings show that diabetes is poorly managed in Bangladesh, especially in men. Among those with diabetes, only 31% were aware of their condition, and 28% were receiving treatment. Only 26% had controlled diabetes mellitus among those who received treatment. Factors independently associated with awareness and treatment were age, sex, hypertension status, and level of education. Factors associated with poor control were secondary education and residing in Rajshahi and Rangpur divisions.

We found that the prevalence of awareness, treatment, and control of diabetes were low. Awareness was lower than the average of South Asian countries (50%) [26], Nepal (65%) [8] and China (49%) [27]. Treatment of diabetes in Bangladesh were also far lower than Nepal (94%) [8] and China (43%). However, the control of diabetes in Bangladesh is higher than Nepal (21%) [8] and China (21%) [27].

This study found awareness and treatment increased with increasing age, which is comparable with other studies conducted in LMICs, including Bangladesh, India, Nepal, and China [7, 8, 11, 28]. A possible explanation is that as people age, they are more likely to have diabetes, be aware they have it, and have the financial resources to obtain appropriate treatment for it. However, recent data indicates diabetes prevalence is also increasing in younger people [29], particularly among educated youth [30]. This change in Bangladesh is consistent with an “Asian phenotype” of diabetes, characterised by an onset of diabetes at a younger age and higher risk even at lower body mass index [31]. Our data suggest that being young does not equate with good diabetes management as awareness and treatment were low in younger people.

We found that people with low level of education were less likely to be aware of, and treated for diabetes than those with higher level of education. [32]. In the Bangladeshi social structure, education is the most important marker of the type of job a person gets. People with lower level of education are most likely to have manual employment involving physical work and have lower health literacy. This is consistent with a low level of awareness and treatment of diabetes [17]. Also, poor control of diabetes is expected among the lower educated respondents, as was found in this study, where a lower likelihood in the control of diabetes was observed in people with secondary education than in people with higher education. This was also in line with previous studies in Bangladesh and India [11, 33].

Though the place of respondents’ residence was not found as a significant factor for awareness and treatment of diabetes, we found poorer control of diabetes in respondents of Rajshahi and Rangpur divisions than in Barishal division. Many factors might contribute to such differences in control of diabetes in these divisions. For instance, Rajshahi and Rangpur divisions are mostly rural and people are mostly engaged with agricultural activity [34]. Consistent with our study, other studies have also found lower awareness, treatment, and control of diabetes among people in these divisions [7, 28], and reported their low level of education and lack of (or difficulties in) access to healthcare facilities [7, 28].

Sex differences in diabetes management were found in this study in which women were more likely to be aware of diabetes and more likely to be treated for diabetes than men. Although statistically not significant, control was also higher in women than men. This is inconsistent with what has been reported in high income countries [35-37] but similar to findings in Bangladesh [11] and China [28]. The better diabetes management in women could be explained by the fact that in the last few decades women are highly exposed to maternal and child health services provided by the community health care workers at the household level. As such women are more likely to discuss a range of health issues, including gestational diabetes, with health workers compared with men [38, 39]. Efforts, particularly in men, are needed to ensure good diabetes management. [40].

Our study shows that people being underweight were less aware of their diabetes condition compared with people having normal weight. One plausible explanation could be the strong association of diabetes with obesity in health promotion literature, reducing the awareness of underweight people to their risk.

The National Guideline for Diabetes Management in Bangladesh focused on raising diabetes awareness and associated factors [41]. These included unhealthy diets, physical activity, and overweight or obesity [41] because of the strength of the evidence of their protective effects [42-44]. However, the roll-out is currently very limited. For instance, public health measures and programs to increase awareness and treatment of diabetes are still restricted to urban centres and are small in scale, mostly organised by the Diabetic Association of Bangladesh [45]. Secondly, public health campaigns alone have not proven effective in preventing diabetes though they can increase awareness [46]. Other innovative approaches including the use of social media and mobile phone text messaging could be a cost effective intervention to improve glycemic control in diabetic patients [47]. This is particularly the case as the government of Bangladesh have adopted information technologies for health in their strategic plans [47].

The implications of our findings are that there needs to be substantial investment in health promotion to raise awareness, and changes in healthcare delivery that address treatment and control of diabetes regardless of socio-demographic status. However, the management of diabetes within the healthcare sector in Bangladesh is challenging for several reasons. The major challenge comes from the facility level, in which maternal and child health and other diseases are prioritised [48]. So far in Bangladesh, the establishment of NCDs’ corners at the Upazila Health Complexes is the only program that the government of Bangladesh has taken to reduce diabetes as well as other NCDs [49]. People with diabetes therefore mostly depend on private healthcare facilities as well as services provided by the Bangladesh Diabetes Federation, a non-profit specialised organisation for managing diabetes. However, their services are mostly located in urban areas, and private healthcare facilities are expensive [48]. Therefore, the management of diabetes is challenging for poor and/rural people due to structural and economic difficulties given that there is no healthcare insurance coverage in Bangladesh.

A limitation of this study was that outcomes data are self-reported. Diet and lifestyle factors are important determinants for diabetes management. However, these data were not available in the dataset; therefore, we could not consider them in our analyses. Moreover, the design of this survey was cross-sectional which limits our capacity to draw casual findings. The major strength is that this is the first study in Bangladesh using a large nationally representative dataset that included adults 18 years and older, suggesting the findings have external validity. Our study generates findings with increased precision because of the use of multilevel mixed-effects Poisson regression that corrects the overestimation of effects size produced by conventional logistic regression employed in cross-sectional studies.

## Conclusion

This study provides evidence of poor management of diabetes in Bangladesh, especially in men. Less than one-third of the people with diabetes were aware of their condition. Just over one-fourth of the people with diabetes were on treatment, and those were treated one-fourth had controlled diabetes. Interventions targeting younger people, in particular men and those with lower education, are urgently needed. The government needs to strengthen diabetes management programs within primary health care and address structural factors including the costs of diabetes care to improve awareness, treatment, and control of diabetes in Bangladesh.

## Data Availability

BDHSs data were collected from the MEASURE DHS. The authors are restricted in sharing or making the dataset publicly available. Interested readers can download this dataset after registering with the MEASURE DHS. Necessary information are available at: http://dhsprogram.com/data/Using-DataSets-for-Analysis.cfm.

## Abbreviations

LMICs: Low- and Lower-Middle-Income Countries
SDGs: Sustainable Development Goals
NCDs: Non-communicable diseases
BDHS: Bangladesh Demographic and Health Survey
PR: Prevalence Ratio
95% CI: 95% Confidence Interval

## Acknowledgement

We acknowledge DHS program of the USA, custodian of the data used in this study, for approving to use their data.

